# Direct and indirect household costs of care of children under 5 years old attending Integrated Management of Childhood Illness consultations at Primary Healthcare Centres in Burkina Faso, Guinea, Mali and Niger: a cross-sectional costing study nested in the longitudinal AIRE project 2021-2022

**DOI:** 10.1101/2024.10.21.24315857

**Authors:** Honorat Agbeci, Richard Bakyono, Amadou Oury Touré, Adama Coulibaly, Zineb Zair, Mactar Niome, Adama Hema, Abdoul-Salam Sawadogo, Sorry Keita, Lucie Peters-Bokol, Gildas Boris Hedible, Désiré Neboua, Sarah Louart, Valérie Zombré, Dieney Fadima Kaba, Amadou Sidibe, Abarry Souleymane Hannatou, Anthony Cousien, Sandrine Busiere, Franck Lamontage, Valéry Ridde, Sophie Desmonde, Valériane Leroy, the AIRE Study Group

## Abstract

**Introduction:** Out-of-pocket payments persist, limiting access to care in Africa. The AIRE project evaluated the implementation of pulse oximetry integrated with Integrated Management of Childhood Illness (IMCI) at Primary Healthcare Centres (PHCs) in Burkina Faso and Niger (total exemption policy) and Mali and Guinea (partial exemption policy). We measured households’ out-of-pocket expenditures for care of children under-5 years and analyzed associated factors.

**Methods:** Between 06/2021-05/2022, five non-severe and five severe cases per PHC among children <5 years attending IMCI consultations (excluding simple non-respiratory cases) in four PHCs per participating country in the AIRE study were selected each month among the children included. Severe IMCI cases were followed-up at 14 days. We collected medical direct costs and non-medical direct and indirect costs. We describe median costs; factors associated with medical direct costs (MDC) were investigated in two-part models for countries with total exemption and general linear model in those with partial exemption.

**Results:** Of the 15,836 children overall, 940 non-severe cases and 745 severe cases were included. The median medical direct costs were USD 0.0, 7.1, 5.0 and 3.6 for non-severe cases, and 1.6, 8.6, 7.4 and 14.4 for severe cases, in Burkina Faso, Guinea, Mali and Niger, respectively. Medicine expenditures were the main MDC item, reaching 79% for non-severe cases and 59% in severe cases. In all countries, the disease severity and the unavailability of prescribed medicines at PHCs or hospital depots were associated to any out-of-pocket payment, and to higher amounts of expenses.

**Conclusion:** With the exception of Burkina Faso and despite free care policies, household out-of-pocket payment remains high for children under-5, particularly for treating severe cases, mainly explained by medicines expenditures. Actions are needed to identify efficient financing systems to ensure regular and adequate delivery of medicines in public health facilities, and to support free healthcare policies.

**Key messages:**

- What is already known on this topic?

- In sub-Saharan Africa, various studies have shown that user fee total or partial exemption policies do not succeed in eliminating or significantly reducing healthcare costs borne by households.
- Few studies have explored household out-of-pocket expenditures for the care of children under 5 years in the West African context.
- What this study adds?

- Despite user-fee exemption policies, household out-of-pocket payment remains high for taking care of children under-5 at primary care and district hospital.
- Most expenditures were associated with purchasing medicines outside of primary healthcare centers and referral hospitals, which is likely the result of stock-outs in public facilities.
- Both the probability of an out-of-pocket expenditure and its amount were associated with the severity of the disease.
- How this study might affect research, practice or policy?

- Our study underlines the importance of further investigations to determine effective funding methods aimed at ensuring a regular and adequate supply of medicines in public healthcare facilities.

**CHEERS Statement:** This study adheres to the Consolidated Health Economic Evaluation Reporting Standards (CHEERS).

## Introduction

Under-5 mortality remains a challenge in sub-Saharan Africa, despite the progress made in child health management worldwide. According to the World Bank, the mortality rate was 73 deaths per 1000 live births in 2021 in this region with infectious diseases remaining the main cause of death (1). Structural factors, such as the limited geographical and financial accessibility of primary care, particularly in rural areas, the lack of adequate diagnostic tools, and shortages of essential medicines contribute to this high mortality (2,3).

Various interventions have been implemented along the continuum of care to improve the accessibility and quality of healthcare services, in order to make progress towards achieving Sustainable Development Goal (SDG) 3 by 2030 (4). In September 1987, many African countries adopted the Bamako initiative (BI) with the support of the World Health Organization (WHO) and the United Nations Children’s Fund (UNICEF), in response to economic crises and difficulties in financing the recurrent costs of primary healthcare (5). This initiative was presented as a major approach to improve maternal and child health by ensuring the availability of essential medicines at the primary healthcare level (6). As part of the BI, direct payments at the point of delivery care were introduced, with local management of revenues by management committees for the purchase of medicines and healthcare provider activities (7,8). However, out-of-pocket payments have been identified in several studies as a major barrier to access to healthcare, exposing households to catastrophic expenditures and a risk of impoverishment (9,10). Since the 2000s, a number of sub-Saharan African countries have implemented selective free care policy, which vary according to the health services covered, the populations benefiting, the level of cost alleviation and the mode of financing. Regarding care policies for children under5 years of age, Burundi and Niger were the first to implement a free care policy, in 2006, followed by Burkina Faso 10 years later (11). Mali and Guinea both introduced a free user fee policy for malaria treatment in 2010 and 2011, in addition to the national programs to fight Human Immunodeficiency Virus (HIV) infection, tuberculosis and malnutrition in these countries (12). Although user fee exemption policies have significantly increased the use of health services, especially among the poorest, out-of-pocket payments in maternal healthcare persist (13–17). Few studies have examined the expenditure incurred by households and their determinants, for children under5 years in the context of partial or total exemption policies (18).

With the goal to reduce under-5 mortality, WHO and UNICEF introduced in 1995 Integrated Management of Childhood Illness (IMCI), a clinical symptom-based algorithm aimed to help health care workers to better diagnose and manage serious illnesses in primary health centers (PHCs) (19–22). However, IMCI does not allow early detection of hypoxemia, which has been identified as a risk factor for death, increasing by 3 the risk of death in children with severe illnesses, respiratory or non-respiratory (23,24). To identify new approaches to reduce hypoxemia-related mortality, the AIRE project (Améliorer l’Identification des détresses Respiratoires de l’Enfant) was implemented to assess the feasibility and added value of the pulse oximeter (PO) integrated with IMCI for the early identification of hypoxemia in PHCs, enabling rapid and appropriate care management in four West African countries in 2021-2022 (25).

The AIRE project offered a unique opportunity to carry out a nested costing study to measure and explore factors associated with household expenditure for children under5 years attending IMCI consultations in PHCs participating in the AIRE project.

## Methods

### Context

The AIRE project, funded by UNITAID, evaluated the implementation of POs integrated with IMCI in Burkina Faso, Guinea, Mali and Niger (25). The healthcare systems in these four countries studied are based on a three-tier pyramid structure. The PHCs at the first level are the theoretical population-entry points to the healthcare system. Based on their condition, patients are referred to their district hospital, with possible transfer to regional hospitals at the secondary level or university hospitals at the third level of the pyramid. The AIRE interventions were implemented in 202 public PHCs, with support to their seven referral hospitals (RHs).

Briefly, IMIC classifies children as green (mild condition requiring simple care at home), yellow (condition manageable at the PHC level, mostly requiring medicines and follow-up) and red (severe condition requiring urgent referral). In AIRE, at all PHCs, all newborns and children aged between 2 and 59 months (except those classified as non-respiratory green) attending IMCI consultations at the PHC level, were eligible for pulse oxygen saturation (SpO2) measurement. Two groups were then defined according to the severity of the disease according to the IMCI integrating PO classification (25). Those classified as "respiratory green" or "yellow", were defined as non-severe cases and those classified as "pink/red" and those with hypoxemia (SpO_2_≤93%) regardless of IMCI were defined as severe cases. All non-severe cases were treated at PHCs on an outpatient basis or underwent observation, followed by treatment at home. Severe cases, in theory, were referred to the RH.

Furthermore, as part of the AIRE project, endowment medicines were provided to the health districts involved in the project for all children treated at the PHCs and their RHs.

### Free care policy

Free care, i.e Total Exemption Policy (TEP) of direct payments was implemented for the care of children under 5 years (consultations, medicines, consumables) in Niger and Burkina Faso in 2006 and 2016, respectively. This means that theoretically, all the available care is provided free of charge at all levels of the health pyramid, regardless of the disease (26). In Niger, facilities receive a lump sum per child cared for, after submitting invoices for free treatment to the Ministry of Health’s free care steering committee who then validates these invoices and orders the disbursement of funds by the Ministry of Finance. In Burkina Faso, facilities are reimbursed the actual costs of care consumed in caring for the child: the government pre-deposits the funds with the health district before the treatment is provided (11,27). This approach was preferred in Burkina Faso in order to avoid reimbursement delays that could affect the proper functioning of the policy (18,28).

In Mali and Guinea, there is a partial exemption policy (PEP) for direct payments, for the care of malaria, malnutrition, tuberculosis and HIV; though medical visit fees persist (29,30). Outside these four diseases, all healthcare services and care are subject to direct payments borne by households (12). Regarding the mode of financing, medicines are directly allocated to health facilities by central medicine purchasing agencies, on the instructions of the government with the support of its partners, who are responsible for paying the bills.

### Study design and population

AIRE research activities were carried out in 16 PHCs (4 PHCs selected per country) and their RHs and included all children aged 0-59 months eligible for PO use, for whom parental consent was given.

Those classified as severe cases were then enrolled in a cohort with a 14-day follow-up (D14). The data collection period ran from June 2021 to May 2022.

The costing sub-study sample was a cross-sectional study carried out in the first five non-severe cases and first five severe cases enrolled in each PHC, over a specific week each month. The selection was carried out over a 12-month period, with a weekly rotation of the selection week over the 4 weeks of the month to account for seasonal variations in diseases (such as malaria during the rainy season and pneumonia during the dry season), as well as variations in households’ ability to pay for care throughout the month.

### Data collection

Costs borne by households are defined in Table 1. Briefly, direct costs were separated into medical (direct payments made by households for health services and care consumed by their child) and non-medical costs (i.e transport to and from the PHC, caregiver accommodation during hospitalisation etc). Expenses related to food and drink and those incurred after death were not considered. Indirect costs represented the loss of income due to the interruption of work or the household’s income-generating activity (IGA) due to the illness. The daily income from field work was USD 1.8 in Burkina Faso, USD 3.6 in Guinea and USD 4.5 in Mali and Niger according to an ad hoc survey of households carried during the period.

**Table 1:** Definition of direct and indirect household costs for the care of children seen in IMCI consultations at the PHC and referral hospital. AIRE project, June 2021 to May 2022.

|  | Direct costs |  | Indirect costs |
| --- | --- | --- | --- |
|  | Medical expenditures | Non-medical expenditures |  |
| **Primary healthcare centre (PHC)** | Primary Healthcare Centre visit | Transport: home to the PHC, purchase of medication and tests, and PHC to home | Loss of income due to interruption of income-generating activity related to illness* |
|  | Medicines |  |  |
|  | Complementary exam |  |  |
|  | All medical and surgical care |  |  |
|  | Medical observation |  |  |
| **Referral hospital (RH)** | Transfer from PHC to hospital | Transport: home to the PHC, purchase of medication and tests, and hospital to home | Loss of income due to interruption of income-generating activity related to illness* |
|  | Hospital admission fees |  |  |
|  | Medicines |  |  |
|  | Complementary exam | Parental housing during hospitalisation |  |
|  | All medical and surgical care, including oxygen therapy | Care for children left at home |  |
|  | Hospitalisation |  |  |

PHC: Primary Healthcare Centre
\* Amount of loss of income was estimated by multiplying the total number of days lost by household members who had an income-generating activity by the daily income from field work in each context. Intangible costs were not considered.

Cost data were collected after discharge from the PHC, outside the facility, on day 5 after the index IMCI consultation for non-severe cases, and during hospital stay then on day 14 for severe cases. Costs were collected based on receipts or, failing that, declarative, in the national currency of each country. Costs were converted into United States Dollar (USD) using the official average exchange rate for 2021 (USD 1 = 9,838.05 Guinean francs and USD 1 =554.53 CFA francs) (31,32). Cost data have not been adjusted or discounted.

Sociodemographic and clinical data, as well as the main diagnosis based on the IMCI diagnosis blocks or the ICD-10 code, were collected (33).

### Analysis

Distributions of characteristics between children selected for the cost study and children not selected and characteristics of all children enrolled in the cost study stratified according to the disease severity, and by country were described and compared using the Pearson’s chi-square test, Wilcoxon signed-ranks test or Student’s t-test, where appropriate, at the 5% significance level.

Qualitative variables were presented as numbers (percentages), while quantitative variables were expressed as mean (standard deviation) or median (Q1: 1st quartile; Q3: 3rd quartile), depending on their distribution.

Costs in 2021 USD were described in median terms (Q1; Q3) for all children by country. To determine the proportion of households with high health expenditure, we used the concept of excessive expenditure rather than catastrophic expenditure due to lack of information on household consumption and/or income (34,35). In each country, households with direct medical costs above a threshold amount determined by the Tukey method (Q3+k*(Q3-Q1), k=1.5)) defined excessive expenditure (36).

Factors associated with costs were investigated in two-part models for Burkina Faso and Niger, where there is TEP, first describing the probability of a household out-of-pocket expenditure with a logistic regression, before identifying associated factors with the costs among those who had health expenditures using an ordinary least squares (OLS) regression model with log transformation of costs (chosen according to the Box-Cox test, p < 0.05 for λ = 0) (37). In Guinea and Mali where only PEP exists and a majority of households experienced out-of-pocket payments, factors associated with the amount of direct payments were investigated in standard generalized linear models (GLM) : the Box-cox test in Guinea (p<0.05 for λ = 0) and Mali (p = 0.993 for λ = 0) and the modified Parks test (Coefficient = 3.3) led us to use a log link and Gaussian distribution in Guinea and inverse Gaussian distribution in Mali. If no payments were made (45 children in Guinea, and 10 in Mali), direct payments were set at USD 0.1 for modelling purposes.

All models were adjusted for socio demographic variables (age, sex and severity of the illness level of education of the head of the household, IGA of the main child‘s caregiver), accessibility to PHC variables (delay since the onset of symptoms, transport means used and length of the transport from home to PHC), structural variables (place of acquisition of prescribed medicines: PHC or RH depot or private pharmacies). The place of medicines acquisition was used as an indicator of their actual availability at PHC or RH depot, assuming that households would prefer to obtain medicines from the PHC depot, as they are provided free of charge in Burkina Faso and Niger or at more affordable prices than in private pharmacies in Guinea and Mali.

All analyses were performed in Stata Version 14.2 software (Stata Corporation, Texas, USA).

### Patient and public involvement

This study was conducted using programmatic data collected routinely with the MOH authorization. Patients were not involved in the analysis plan or result interpretation. Patients did not contribute to the writing or editing of this manuscript.

## Results

### Study sample

Between June 2021 and May 2022, of the 39,360 children <5 years attending IMCI consultations in the 16 AIRE research PHCs, 15,836 were included in the AIRE main analyses in Burkina Faso, Guinea, Mali and Niger (Figure 1). Detailed description of children enrolled is provided elsewhere (25). At PHC level 941(6.8%) of the 13,838 non-severe IMCI cases identified, and 757 (37.9%) of the 1,998 severe cases were selected for the cost study. After exclusion of missing and loss to follow-up data, 940 (99.8%) non-severe cases and 745 (98.4%) severe cases were analyzed. A comparison of individual and family characteristics between children selected in the cost sample and the children not selected according to illness severity and country showed an independent distribution in the two groups, with the exception of Mali. The proportion of children whose principal caregiver has an IGA was significantly higher in those included in our study than those not selected, for both severe and non-severe cases in Mali (Appendix 1).

**Figure 1:**
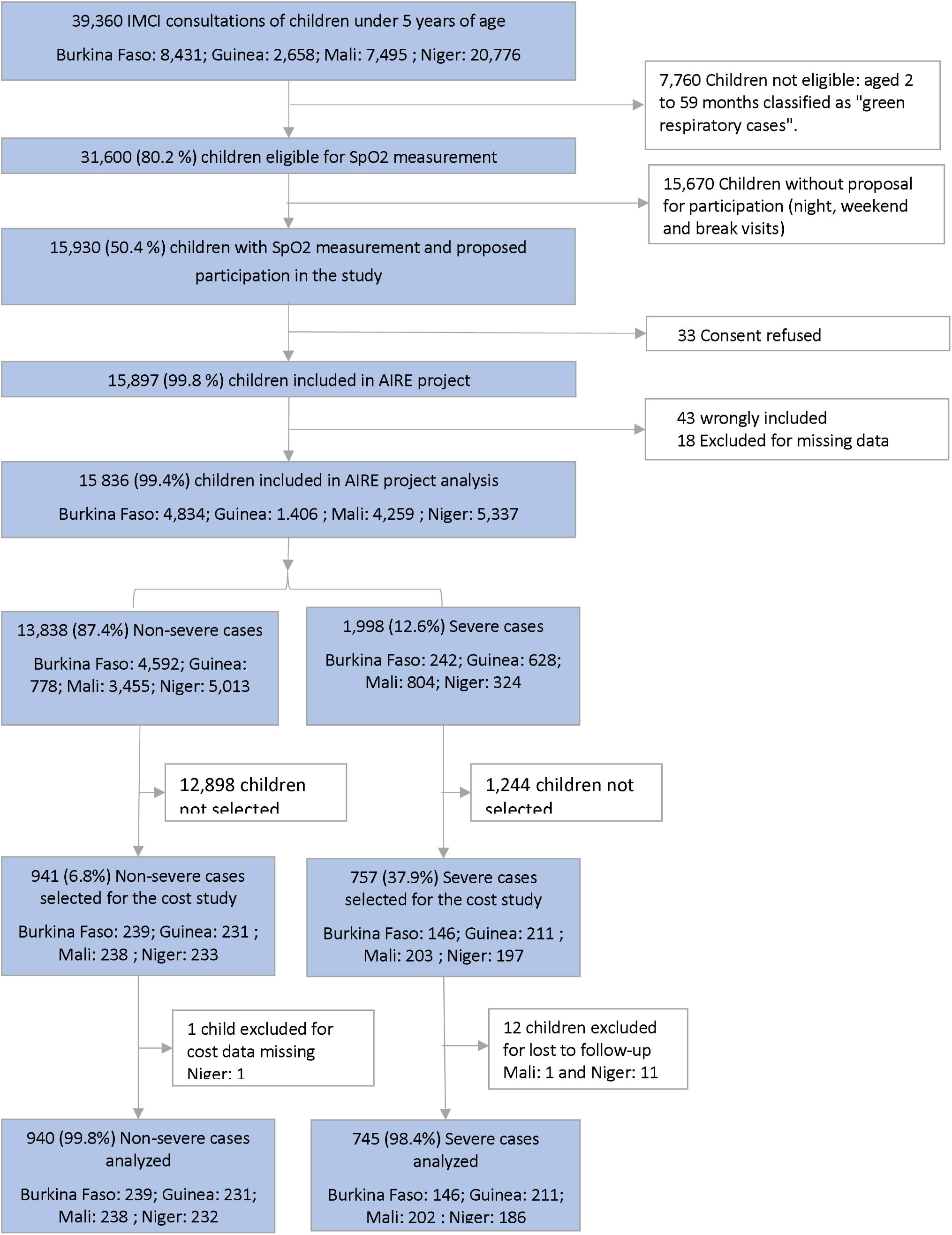
Flow chart of children selected and included in the cost analysis in the AIRE project, June 2021 to May 2022.

Characteristics of children included are described in table 2. Median age ranged from 13 months [8-26] in Niger to 26 months [3-43] in Burkina Faso. In more than half to 80% of households, the head of household had never attended school, and with the exception of Guinea, only less than a third of child’s main caregivers has an income-generating activity. Overall, IMCI visits to PHC were mostly carried out later than two days after the onset of symptoms, with higher proportions for severe cases compared to non-severe cases. The transport duration from home to the PHC was less than 30 minutes for most households. Overall, malaria was the most frequent main diagnosis retained, ranging from 31% to 50% for non-severe cases and from 48% to 90% for severe cases, followed by respiratory diseases and malnutrition. Among severe cases, in Niger, 74% were referred to RH, this was much higher than in other countries (29% in Burkina Faso,13% in Mali and 19% in Guinea). In Burkina Faso, Guinea and Mali, medicines were purchased within the PHC or hospital depot, subsidised according to country-specific payment exemption policies. In Niger, 86% of non-severe cases and 54% of severe cases, purchased medicines outside of PHCs or hospitals.

**Table 2:** Characteristics of children included at PHCs in the cost analysis according to exemption user fees policy, severity of the disease and country, AIRE project, June 2021-May 2022.

|  | Non-severe cases |  |  |  |  | Severe cases |  |  |  |  |
| --- | --- | --- | --- | --- | --- | --- | --- | --- | --- | --- |
|  | Total exemption policy |  | Partial exemption policy |  | *p-value* | Total exemption policy |  | Partial exemption policy |  | *p-value* |
|  | Burkina Faso<br>(N=239) | Niger<br>(N=232) | Guinea<br>(N=231) | Mali<br>(N=238) |  | Burkina Faso<br>(N=146) | Niger<br>(N=186) | Guinea<br>(N=211) | Mali<br>(N=202) |  |
| **Median age, months [q1 - q3]** | 18 [7-36] | 13 [8-26] | 19 [8-34] | 17 [8-36] | 0.028 | 26 [3-43] | 16 [9-24] | 24 [11-36] | 19 [7-36] | 0.002 |
| **Male sex, n (%)** | 132 (55) | 124 (53) | 114 (49) | 138 (58) | 0.295 | 87 (60) | 101 (54) | 113 (54) | 104 (51) | 0.505 |
| **Education level of the head of household, n (%)** |  |  |  |  | <0.001 |  |  |  |  | <0.001 |
| None | 192 (80) | 133 (57) | 120 (52) | 152 (64) |  | 123 (84) | 138 (74) | 111 (53) | 140 (69) |  |
| Primary level and up | 47 (20) | 99 (43) | 111 (48) | 86 (36) |  | 23 (16) | 48 (26) | 100 (47) | 62 (31) |  |
| **Income-generating activity of the child's caregiver, n (%)** | 61 (26) | 57 (25) | 126 (55) | 76 (32) | <0.001 | 24 (16) | 26 (14) | 107 (51) | 56 (28) | <0.001 |
| **Time since onset of symptoms ≥2 days, n (%)** | 135 (56) | 128 (55) | 195 (84) | 160 (67) | <0.001 | 80 (55) | 153 (82) | 182 (86) | 145 (72) | <0.001 |
| **Means of transport to PHC, n (%)** |  |  |  |  | <0.001 |  |  |  |  | <0.001 |
| Foot/Cart/Bike | 192 (81) | 186 (80) | 84 (37) | 64 (27) |  | 85 (59) | 117 (63) | 71 (34) | 36 (18) |  |
| Taxi: car/Motorcycle/Bus | 46 (19) | 46 (20) | 145 (59) | 172 (73) |  | 60 (41) | 68 (37) | 140 (60) | 161 (82) |  |
| **Transport duration from home to PHC ≥30 minutes, n (%)** | 131 (55) | 58 (25) | 50 (22) | 49 (21) | 0.004 | 15 (10) | 79 (42) | 65 (31) | 54 (27) | <0.001 |
| **Main diagnosis of child illness\*, n (%)** |  |  |  |  | <0.001 |  |  |  |  | <0.001 |
| Malaria | 87 (36) | 73 (31) | 116 (50) | 81 (34) |  | 82 (56) | 79 (48) | 189 (90) | 133 (68) |  |
| Respiratory disease | 64 (27) | 60 (25) | 66 (29) | 54 (23) |  | 11 (8) | 38 (23) | 5 (2) | 26 (13) |  |
| Malnutrition | 8 (3) | 30 (13) | 19 (8) | 39 (16) |  | 11 (8) | 39 (23) | 3 (1) | 10 (5) |  |
| **Place of care management, n (%)** |  |  |  |  |  |  |  |  |  | <0.001 |
| At Primary Health Center (PHC) | 239 (100) | 232 (100) | 231 (100) | 238 (100) | NA | 103 (71) | 50 (27) | 192 (91) | 168 (83) |  |
| At PHC, then Referral Hospital (RH) | NA | NA | NA | NA |  | 43 (29) | 136 (73) | 19 (9) | 34 (17) |  |
| **Place of purchase of medicines n (%)** |  |  |  |  | <0.001 |  |  |  |  | <0.001 |
| PHC or RH | 169 (71) | 33 (14) | 194 (84) | 214 (90) |  | 131 (90) | 85 (46) | 170 (81) | 185 (92) |  |
| At least one purchased outside of PHC or RH | 70 (29) | 199 (86) | 37 (16) | 24 (10) |  | 15 (10) | 101 (54) | 41 (19) | 17 (8) |  |
| **Death at D14** | NA* | NA* | NA* | NA* |  | 8 (5) | 19 (10) | 1 (0) | 15 (7) | <0.001 |

\* IMCI diagnosis for non-severe cases, or ICD-10 code for severe cases ; NA: Not applicable ; NA\*: Not available.

### Costs

The median [Q1-Q3] medical direct costs (MDC) for the management of non-severe cases at the PCH were USD 0 [0 - 1.4], 3.6 [1.9 - 5.6], 5.0 [3.8 - 6.7] and 7.1 [5.1 - 9.1] in Burkina Faso, Niger, Mali and Guinea, respectively (Table 3). Among non-severe cases, the main item for household health expenditure was the purchase of medicines ranging from 79% of total MDC in Guinea to 100% in Burkina Faso. In addition, consultation fees at the PHC level represented 21%, 12% and 1% in Mali, Guinea and Niger, respectively (Appendix 2).

**Table 3.** Direct and indirect costs for households (median cost for all children and mean cost for children whose households have incurred expenses) in 2021 US dollars per child under 5 seen in IMCI consultations at PHCs and referral hospital, according to case severity, exemption policy and country, AIRE project, June 2021-May 2022.

|  | Total exemption policy |  |  |  |  |  | Partial exemption policy |  |  |  |  |  |
| --- | --- | --- | --- | --- | --- | --- | --- | --- | --- | --- | --- | --- |
|  | Burkina Faso |  |  | Niger |  |  | Guinea |  |  | Mali |  |  |
|  | Non-severe cases (N=239) | Severe cases (N=146) | p-value | Non-severe cases (N=232) | Severe cases (N=186) | p-value | Non-severe cases (N=231) | Severe cases (N=211) | p-value | Non-severe cases (N=238) | Severe cases (N=202) | p-value |
| **Medical direct cost** |  |  |  |  |  |  |  |  |  |  |  |  |
| **Median cost [q1-q3] of all children included** | 0 [0 - 1.4] | 1.6 [0 - 6.0] | <0.001 | 3.6 [1.9 - 5.6] | 14.4 [6.7 - 22.7] | <0.001 | 7.1 [5.1 - 9.1] | 8.6 [3.1 - 13.2] | <0.001 | 5.0 [3.8 - 6.7] | 7.4 [0.9 - 13.9] | <0.001 |
| PHC visit / Hospital admission fees | 0 [0 - 0] | 0 [0 - 0] |  | 0 [0 - 0] | 0 [0 - 0] |  | 0.5 [0.5 - 1.0] | 1.1 [0 - 1.1] |  | 0.9 [0.9 - 0.9] | 0.9 [0.9 - 0.9] |  |
| Medicines prescribed at PHC and/or hospital | 0 [0 - 1.4] | 0 [0 - 5.2] |  | 3.6 [1.9 - 5.5] | 13.6 [6.3 - 21.3] |  | 6.1 [4.6 - 7.6] | 7.1 [0 - 10.7] |  | 4.1 [2.9 - 5.6] | 5.9 [0 - 10.8] |  |
| Complementary exams | 0 [0 - 0] | 0 [0 - 0] |  | 0 [0 - 0] | 0 [0 - 0] |  | 0 [0 - 0] | 1.1 [0 - 2.0] |  | 0 [0 - 0] | 0 [0 - 0] |  |
| Transfer from PHC to hospital | NA | 0 [0 - 1.1] |  | NA | 0 [0 - 1.8] |  | NA | 0 [0 - 0] |  | NA | 0 [0 - 0] |  |
| Other of direct medical cost item* | 0 [0 - 0] | 0 [0 - 0] |  | 0 [0 - 0] | 0 [0 - 0] |  | 0 [0 - 0] | 0 [0 - 0] |  | 0 [0 - 0] | 0 [0 - 0] |  |
| **Mean (SD) cost:** |  |  |  |  |  |  |  |  |  |  |  |  |
| All children included | 0.9 (2.1) | 5.6 (10.3) |  | 4.0 (2.9) | 18.8 (16.8) |  | 7.3 (3.7) | 12.1 (14.3) |  | 5.5 (3.0) | 12.2 (18.4) |  |
| Children with households expenditures (cost>0) | 3.4 (2.8) | 9.9 (12.1) |  | 4.6 (2.7) | 20.2 (16.6) |  | 7.5 (3.6) | 14.9 (14.5) |  | 5.5 (2.9) | 12.8 (18.6) |  |
| **Non-Medical direct cost** |  |  |  |  |  |  |  |  |  |  |  |  |
| **Median cost [q1-q3] of all children included** | 0 [0 - 0] | 0.6 [0 - 1.8] | <0.001 | 0 [0 - 0] | 1.4 [0 - 2.7] | <0.001 | 0 [0 - 0] | 1.0 [0 - 2.0] | <0.001 | 0 [0 - 0] | 1.2 [0.7 - 2.2] | <0.001 |
| Transport: home to the PHC, purchase of medication and tests, and PHC to home | 0 [0 - 0] | 0.6 [0 - 1.8] |  | 0 [0 - 0] | 1.4 [0 - 2.7] |  | 0 [0 - 0] | 1.0 [0 - 2.0] |  | 0 [0 - 0] | 1.3 [0.7 - 2.2] |  |
| Other direct non-medical cost items** | 0 [0 - 0] | 0 [0 - 0] |  | 0 [0 - 0] | 0 [0 - 0] |  | 0 [0 - 0] | 0 [0 - 0] |  | 0 [0 - 0] | 0 [0 - 0] |  |
| **Mean (SD) cost:** |  |  |  |  |  |  |  |  |  |  |  |  |
| All children included | 0.0 (0.2) | 2.1 (5.5) |  | 0.0 (0.2) | 2.4 (3.1) |  | 0.1 (0.6) | 1.3 (1.3) |  | 0.0 (0.0) | 2.2 (7.2) |  |
| Children with households expenditures (cost>0) | 1.2 (0.4) | 3.7 (6.9) |  | 1.5 (0.6) | 3.2 (3.3) |  | 1.9 (1.0) | 1.8 (1.2) |  | 0 (0.0) | 2.6 (7.8) |  |
| **Indirect cost** |  |  |  |  |  |  |  |  |  |  |  |  |
| **Median cost [q1-q3] of all children included** | 0 [0 - 0] | 0 [0 - 3.6] | <0.001 | 0 [0 - 0] | 0 [0 - 0] | 0.039 | 0 [0 - 0] | 0 [0 - 3.6] | <0.001 | 0 [0 - 0] | 4.5 [0 - 9.0] | <0.001 |
| **Mean (SD) cost:** |  |  |  |  |  |  |  |  |  |  |  |  |
| All children included | 0.0 (0.0) | 3.2 (5.5) |  | 0.0 (0.0) | 0.7 (5.9) |  | 1.1 (3.2) | 3.4 (5.4) |  | 0.6 (1.6) | 6.5 (11.5) |  |
| Children with households expenditures (cost>0) | 0.0 (0.0) | 8.0 (6.2) |  | 0.0 (0.0) | 29.7 (22.9) |  | 6.1 (5.4) | 7.9 (5.7) |  | 4.8 (1.1) | 12.7 (13.4) |  |

NA: Not applicable
\* Medical and surgical care, including oxygen therapy and Medical observation / Hospitalisation
\*\* Parental housing during hospitalisation and Care for children left at home

The MDC for the management of severe cases, wherever their place of management (PHC and/or RH) ranged from USD 1.6 [0 - 6.0], 7.4 [0.9 - 13.9], 8.6 [3.1 - 13.2] to 14.4 [6.7 - 22.7] in Burkina Faso, Mali, Guinea and Niger, respectively (Table 3). Similarly to non-severe cases, the purchase of medicines was the main item of expenditure, representing 75%, 90%, 73% and 59% of total MDC in Burkina Faso, Niger, Mali and Guinea, respectively. The second item of expenditure depends on the exemption policy. In countries with TEP, it was the cost of transferring children from the PHCs to the RH, reaching 23% in Burkina Faso (Appendix 2). Among households who paid for the transfer from PHC to RH, the mean expenditure was USD 3.0 in Burkina Faso, USD 6.1 in Niger USD 2.7 (Appendix 3). In countries with PEP, it was related to PHC visit and hospital admission fees (32%) in Mali, and to exam cost (16%) in Guinea (Appendix 2).

Overall, in Burkina Faso and Niger where TEP exists, 38% (148/385) and 89% (374/418) respectively of the households made at least one direct payment for their child’s care. Of these, the mean MDC was USD 7.0 in Burkina Faso and USD 11.8 in Niger. In Mali and Guinea with PEP, the mean cost was USD 8.8 and 10.6 respectively. For severe cases, MDC mean costs ranged from USD 9.9 in Burkina Faso to USD 20.2 in Niger (Appendix 4). MDC costs were defined as excessive for 26.4%, 10.2%, 7.3%, and 11.9% of the households in Burkina Faso, Niger, Guinea and Mali, respectively (Appendix 5).

For all countries, the median non-medical direct cost was USD 0 [0-0] for non-severe cases and ranged from USD 0.6 [0 - 1.8] in Burkina Faso to 1.4 [0 - 2.7] in Niger for severe cases. Non-medical direct costs were mainly transport expenses from home to PHC (Table 3).

Among severe cases, in Burkina Faso, Guinea and Mali, over a third of households familly members stopped working to care for their child; in Niger, this proportion was 2.7%. The average estimated loss of income per episode of serious illness ranged from USD 7.9 in Guinea to USD 29.7 in Niger (Appendix 4).

Medical direct costs, non-medical direct costs and indirect costs were significantly higher for severe cases compared to non-severe cases in all of the countries (Table 3).

### Factors associated with direct payments

Table 4 (4a-4d) presents factors associated with direct payments for the four countries separately, in adjusted analyses.

**Table 4:**
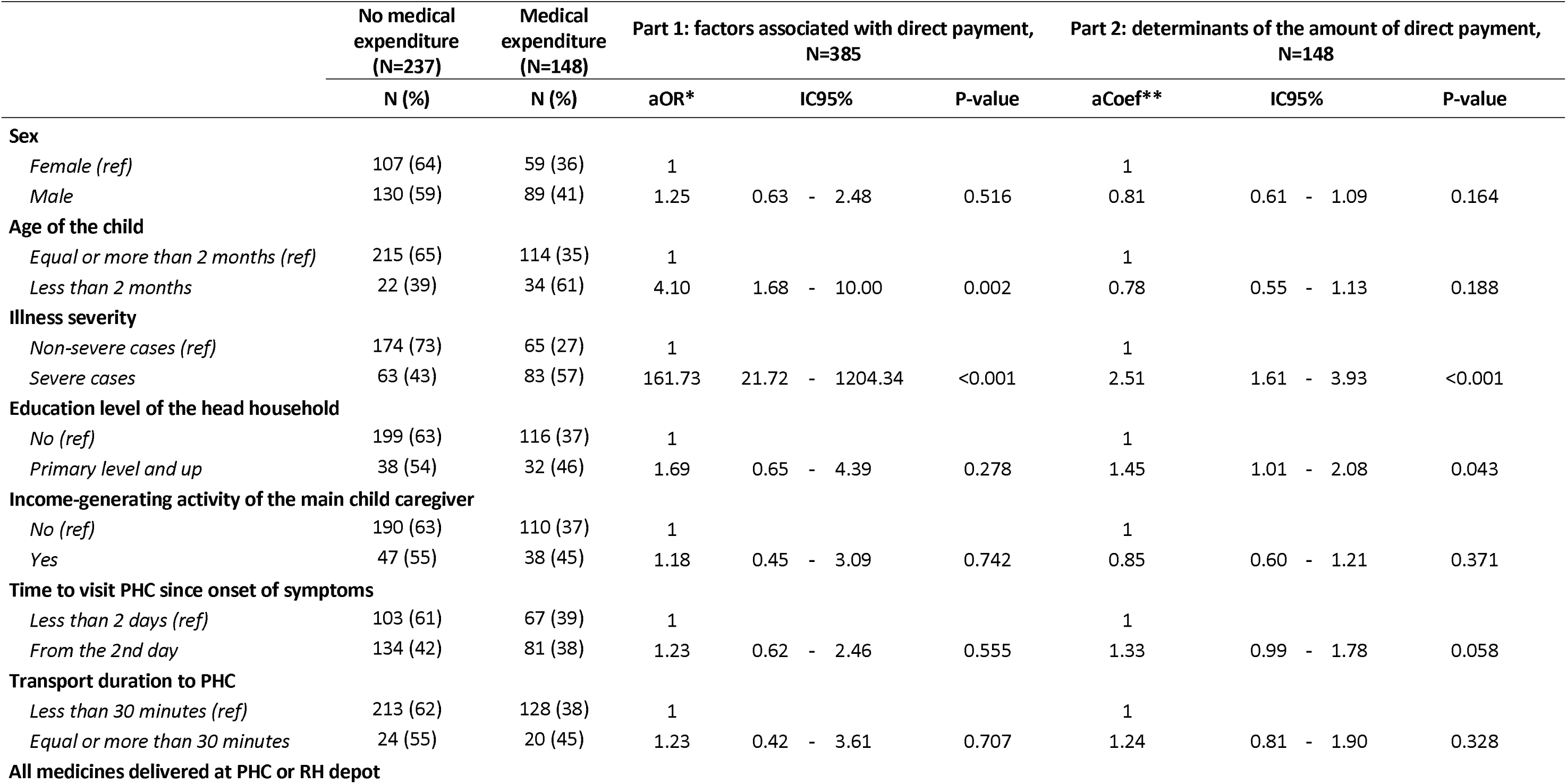
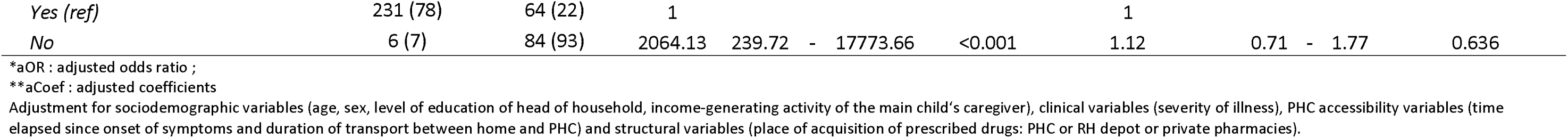
Factors associated with direct payments for the care of children under-5 attending IMCI consultations in Burkina Faso (table 4a) and Niger (table 4b), Guinea (table 4c) and Mali (table 4d). AIRE project, June 2021 to May 2022. **Table 4a:** Factors associated with direct payments in Burkina Faso using an adjusted two-part model (Part 1: logistic regression, Part 2: Ordinary Least Squares with log transformation). N=385

**Table 4b:**
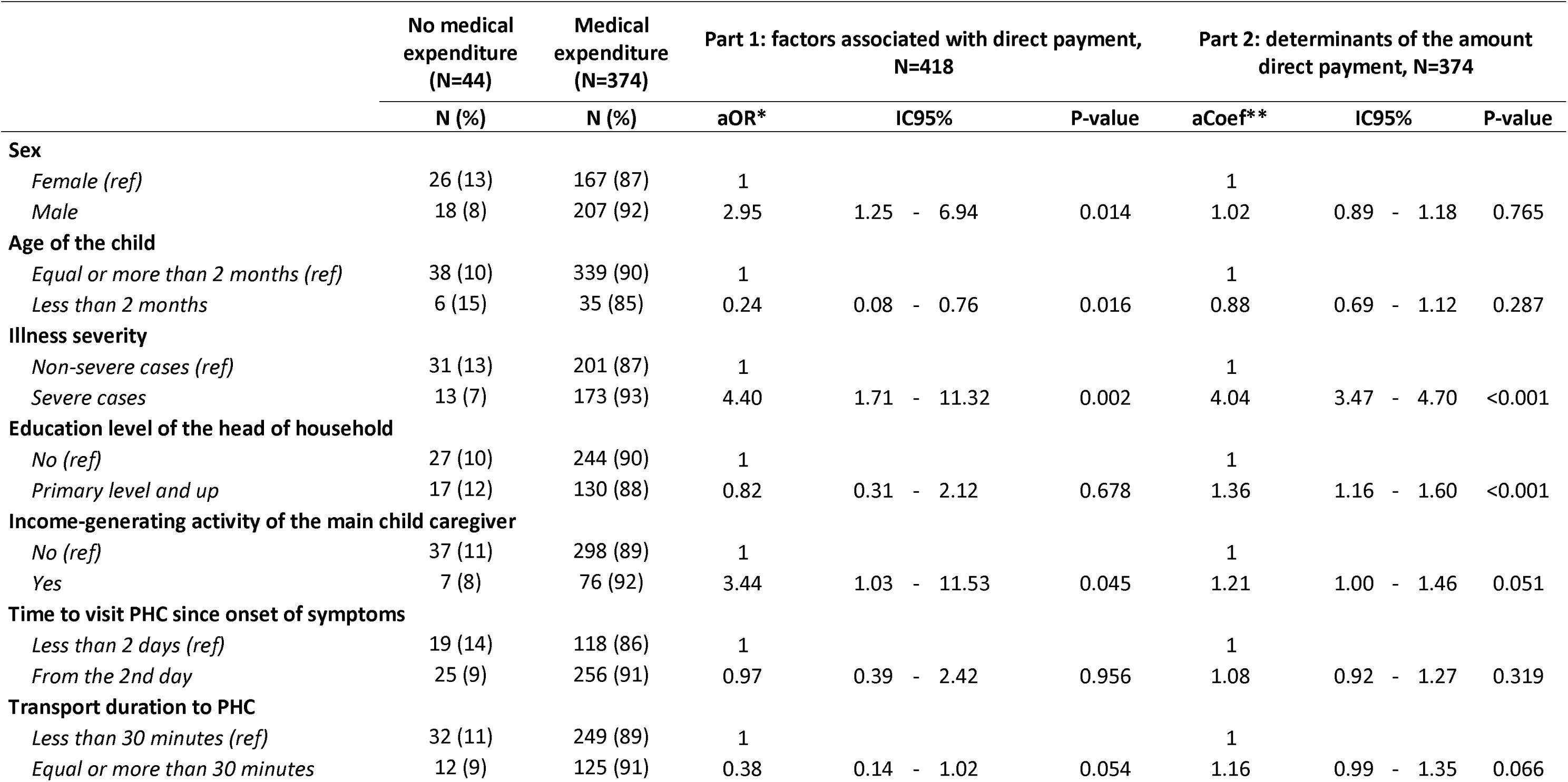
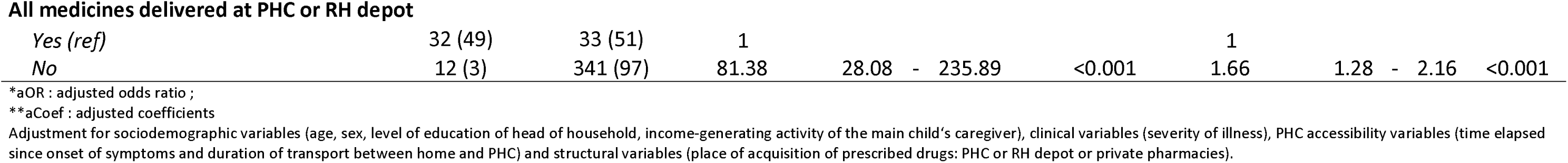
Factors associated with direct payments in Niger using an adjusted two-part model (Part 1: logistic regression, Part 2: Ordinar y Least Squares with log transformation) N=418.

**Table 4c:** Factors associated with direct payments in Guinea using an adjusted Generalized linear model with a log link and gaussian distribution. N=442.

|  | aCoef* | IC95% |  |  | P-value |
| --- | --- | --- | --- | --- | --- |
| **Sex** |  |  |  |  |  |
| Female (ref) | 1 |  |  |  |  |
| Male | 1.25 | 1.07 | - | 1.46 | 0.006 |
| **Age of the child** |  |  |  |  |  |
| Equal or more than 2 months (ref) | 1 |  |  |  |  |
| Less than 2 months | 0.48 | 0.24 | - | 0.94 | 0.032 |
| **Illness severity** |  |  |  |  |  |
| Non-severe cases (ref) | 1 |  |  |  |  |
| Severe cases | 2.00 | 1.62 | - | 2.45 | <0.001 |
| **Education level of the head of household** |  |  |  |  |  |
| No (ref) | 1 |  |  |  |  |
| Primary level and up | 0.70 | 0.59 | - | 0.82 | <0.001 |
| **Income-generating activity of the main child caregiver** |  |  |  |  |  |
| No (ref) | 1 |  |  |  |  |
| Yes | 1.05 | 0.89 | - | 1.23 | 0.561 |
| **Time to visit PHC since onset of symptoms** |  |  |  |  |  |
| Less than 2 days (ref) | 1 |  |  |  |  |
| From the 2nd day | 1.11 | 0.84 | - | 1.47 | 0.468 |
| **Transport duration to PHC** |  |  |  |  |  |
| Less than 30 minutes (ref) | 1 |  |  |  |  |
| Equal or more than 30 minutes | 0.97 | 0.81 | - | 1.15 | 0.699 |
| **All medicines delivered at PHC or RH depot** |  |  |  |  |  |
| Yes (ref) | 1 |  |  |  |  |
| No | 2.97 | 2.49 | - | 3.54 | <0.001 |

\*aCoef : adjusted coefficients
Adjustment for sociodemographic variables (age, sex, level of education of head of household, income-generating activity of the main child's caregiver), clinical variables (severity of illness), PHC accessibility variables (time elapsed since onset of symptoms and duration of transport between home and PHC) and structural variables (place of acquisition of prescribed drugs: PHC or RH depot or private pharmacies).

**Table 4d:** Factors associated with direct payments in Mali using an adjusted Generalized linear model with log link and inverse gaussian distribution N=440.

|  | aCoef | IC95% |  |  | P-value |
| --- | --- | --- | --- | --- | --- |
| **Sex** |  |  |  |  |  |
| Female (ref) | 1 |  |  |  |  |
| Male | 1.03 | 0.87 | - | 1.23 | 0.719 |
| **Age of the child** |  |  |  |  |  |
| Equal or more than 2 months (ref) | 1 |  |  |  |  |
| Less than 2 months | 0.97 | 0.72 | - | 1.31 | 0.853 |
| **Illness severity** |  |  |  |  |  |
| Non-severe cases (ref) | 1 |  |  |  |  |
| Severe cases | 2.19 | 1.79 | - | 2.68 | <0.001 |
| **Education level of the head of household** |  |  |  |  |  |
| No (ref) | 1 |  |  |  |  |
| Primary level and up | 1.01 | 0.83 | - | 1.22 | 0.944 |
| **Income-generating activity of the main child caregiver** |  |  |  |  |  |
| No (ref) | 1 |  |  |  |  |
| Yes | 0.87 | 0.72 | - | 1.04 | 0.125 |
| **Time to visit PHC since onset of symptoms** |  |  |  |  |  |
| Less than 2 days (ref) | 1 |  |  |  |  |
| From the 2nd day | 1.00 | 0.82 | - | 1.21 | 0.961 |
| **Transport duration to PHC** |  |  |  |  |  |
| Less than 30 minutes (ref) | 1 |  |  |  |  |
| Equal or more than 30 minutes | 0.98 | 0.79 | - | 1.22 | 0.867 |
| **All medicines delivered at PHC or RH depot** |  |  |  |  |  |
| Yes (ref) | 1 |  |  |  |  |
| No | 1.44 | 1.05 | - | 1.98 | 0.024 |

\*aCoef : adjusted coefficients
Adjustment for sociodemographic variables (age, sex, level of education of head of household, income-generating activity of the main child's caregiver), clinical variables (severity of illness), PHC accessibility variables (time elapsed since onset of symptoms and duration of transport between home and PHC) and structural variables (place of acquisition of prescribed drugs: PHC or RH depot or private pharmacies).

In Burkina Faso and Niger, where there are TEPs, the strongest independent factors for any out-of-pocket payments were the severity of the disease and the purchase of at least one prescribed medicine outside the PHC or RH depot. In Burkina Faso, families of children < 2 months were four times as likely to make out-of-pocket payments compared to older children 2-59 months, whereas in Niger, families of younger children were less likely to make out-of-pocket payments compared to 2-59 month-olds (aOR = 0.24; 95% CI: 0.08 - 0.76) Furthermore, in Niger we found that out-of-pocket payments were more likely for boys compared to girls (aOR=2.95; 95% CI: 1.25-6.94) and when the principal caregiver had an IGA (aOR=3.4; 95% CI: 1.03-11.53) (Table 4b).

In all countries, regardless of exemption policies, expenses were significantly higher among severe cases compared to non-severe cases. Other determinants varied per country. Among those who had expenses, these were higher where the household head had a primary school or higher education compared to no education in Burkina Faso (acoefficient=1.45; 95%CI: 1.01-2.08) and in Niger (acoefficient=1.36; 95% CI: 1.16-1.60). In Niger, Mali and Guinea, we found that purchasing medicine outside the PHC/hospital depot was associated with significantly higher expenses. In addition, in Guinea, expenses were significantly higher in boys compared to girls (acoefficient: 0.48; 95% CI: 0.24-0.94) and in older children aged 2-59 months compared to < 2 months (acoefficient: 0.97; 95% CI: 0.72-1.31) in Mali.

## Discussion

Our study provides original findings on estimating the household costs of care and associated factors for IMCI children under 5 seeking care at PHCs according to their disease severity, and separately in four West African countries. We report several findings. First, households are subject to out-of-pocket payments in all the countries, regardless of any kind of exemption policy. Second, we found that these payments were mostly for purchasing medicines outside of PHCs and RHs and when the case was severe. Third, in multivariate analyses, we found that determinants of out-of-pocket payments and the amount of these expenses varied by setting, but overall, purchasing medicines outside of the PHC or RH and case severity were strongly associated with both having an out-of-pocket payment in settings where there is TEP and with the amount of these payments in all countries except in Burkina Faso where purchasing medicines outside of the PHC or RH were not associated to the amount.

Overall, the medical direct costs borne by households for the management of any episode of illness were the lowest in Burkina Faso, but highest for the care management of severe cases in Niger, reaching up to USD 14.4, despite the total exemption policies for the health care of children under 5 implemented in Burkina Faso and Niger. Furthermore, in countries with PEP, out-of-pocket payments amounted reached to USD 7.4 and USD 8.6, respectively in Mali and in Guinea, despite malaria being the main diagnosis in more than two-thirds of the severe cases, and total exemption of direct payments for malaria management. In addition to MDC, we also report high transport costs to and from the PHC from home and loss of income associated with severe cases. In our study setting, the minimum wage varies from USD 49 to 72 in the formal sector and 13.8% and 50.6% of the population live below the extreme poverty threshold (USD 2.15 per day) (38–42). These significant non-medical and medical direct costs associated with the care of a child and borne by household do not allow PEP or TEP alone to protect households from the poverty trap. Our results highlight the insufficiency of current free care policies aimed at eliminating or reducing household direct payments to improve affordability of primary care for children under 5. This can lead to delayed access to care: we found that a high proportion of children presented at their PHC more than two days after the onset of symptoms.

Regardless of the case severity and country, nearly all MDCs were explained by the purchase of medicines in private pharmacies, outside of the PHCs and RHs. These purchases are most likely a result of the lack of availability and stock outs of medicines in the public PHC and RH depots. Furthermore, we found that MDCs were, greater for severe cases and in some settings, such as Burkina Faso, that out-of-pockets payments were associated with young age suggesting that medicines stock-outs may also be related to the lack of specific galenic forms suitable for the youngest children, including children under 2 months of age. Additionally, we hypothesise that essential medicines supposed to be delivered to public structures face frequent stock-outs according to different mechanisms depending on payment policies. In Burkina Faso and Niger where there are TEPs, medicine stock-outs are likely attributed to delays in reimbursing invoices issued by public health facilities for the care delivered free of charge to beneficiaries, resulting in an irregular supply of medicines used by these beneficiaries at PHC and HRs pharmacy depots. In Niger, a study reported in 2009 that reimbursement delays reached up to 9 months in one health district (43). In addition, the lump sum given by the government is not sufficient to adequately reimburse health structures in Niger (27). Since the introduction of the TEP in Niger in 2006, the delay in reimbursing invoices issued by health facilities has amounted to 25 billion CFA francs in 2015, according to a study by Niger’s national statistics institute (44). This financial shortfall likely leads to frequent stock-outs, which could persist over the long term. In Niger’s PHCs, only 14 % of non-severe cases received all the medicines prescribed free of charge, compared with 71% in Burkina Faso.

In hospitals, these proportions were 46% and 90% respectively. Of note, in Burkina Faso, since the implementation of the TEP in 2016, out of a total of 32 billion CFA francs in reimbursement requests, 26 billion had been reimbursed at the end of 2019 (45). In Mali and Guinea who have implemented PEPs, and for whom we make the same observations, stock-outs may be the result of insufficient or delayed delivery from the public pharmaceutical depots (30,46). The high amounts of direct payments may be also explained by inappropriate medicine prescriptions, particularly antibiotics overprescription. This is common to all four countries, and according to the WHO, more than 40% of antibiotic prescriptions in primary healthcare in low and middle-income countries are inappropriate (47). In the AIRE research, as investigated elsewhere, we found high proportions of antibiotic prescriptions among children classified as simple IMCI cases in all countries, even though no antibiotic therapy is recommended for the management of these cases (48). We also hypothesise that medicines prescribed were not necessarily those on the list of essential medicines which are supposed to be less costly and more importantly, available in PHCs and RHs.

Out of all four countries, out-of-pocket payments for the management of severe cases were the highest in Niger, despite PEP. This difference can be explained by a higher referral rate to RHs (73% vs 29% in other countries), which leads to additional transfer costs to RH, where care management is more specialised and potentially more expensive. This high referral rate to RH in Niger could be explained by the geographic proximity of the PHCs and the RH particularly in Niamey. These high costs are 90% related to medicine purchases outside of the RH, which is extremely concerning in terms of availability. Furthermore we found that in Niger, care of children < 2 months were less likely to induce expenses. Unfortunately we have data on the outcome only for severe cases, but we hypothesise that these younger may not have received adequate care due to delayed visits to the PHC (82%) and the lack of medicines in appropriate galenic forms and dosages, which led to early deaths resulting in the absence of MDC. There is an urgent need to improve financing modalities to ensure a regular and adequate supply of medicines at all levels of the health pyramid. We found that sex also played a role in out-of-pockets costs. In Niger, an expenditure was up to 3 times more likely in males compared to females.In Guinea out-of-pockets costs were significantly higher in males compared to females. This may be related to socio-cultural households’ preference for male children in Africa, sometimes also expressed by women, as reported in health surveys carried out in 29 African countries (49). This will need further investigation regarding the mortality outcomes of these children.

Our results are in line with previous studies conducted in West Africa and other African settings such as Tanzania which have shown that exemption policies do not make it possible to eliminate direct payments (17,18,43,50–52). Several studies have also shown that the purchasing of medicines outside of the public pharmacy depots, due to stock-outs, was the main determinant of direct payments (35,53). Our results for Burkina Faso are consistent with those of a study carried out there one year after the implementation of the exemption policy, showing that 31% of households had made direct payments, with the following determinants: young age (0 to 6 months) and the severity of the disease. (18).

This study presents several limitations. First, MDC may be underestimated in these countries due to the medicines being supplied as part of the AIRE project, even though these supplies did not specifically target the children included in the project. In some cases, it may have been overestimated. Our results showed that direct payments were higher for children whose head of household had a primary or higher level of education or whose main caregiver had an IGA. Expenditure not incurred due to insufficient resources has not been taken into account. In Mali, the proportion of non-severe and severe cases whose principal caregiver has an IGA was significantly higher in children included in cost study. Non-serious cases were also significantly higher in Burkina Faso. Second, data on household income or consumption were not collected to determine the proportion of households experiencing catastrophic expenditure in each country, but these proportions were obtained using the excessive out-of-pocket expenditure. Finally, it does not provide information on the strategies adopted by households to cope with these expenses.

Nevertheless, our study is one of the first to attempt to measure the direct and indirect costs of caring for children under 5 to households making a significant contribution to the very limited scientific literature on health economic evaluation in these contexts. It thus provides an unique opportunity to compare the implementation of health policies at each national level, using standardised data allowing to better understand obstacles and levers of the success of an exemption policy in sub-Saharan Africa. By reporting harmonised data spent of medical direct costs, mostly based on payment receipts provided by the families, distantly from the influence of the healthcare staff and as close as possible to the health event, between D1 and D5 for non-severe cases and during hospitalisation for severe cases, then at D14 for health expenditure incurred after hospital discharge, we though the results of our study are accurate enough.

## Conclusion

The results of this study, carried out as part of the AIRE project, highlight the persistence of out-of-pocket payments, despite total or partial exemption policies for children under 5. These expenses remain high, particularly for the treatment of severe cases that need to be hospitalised, leading to delayed seeking of care. The probability of an out-of-pocket payment and its expense was strongly associated with the purchase of medicines outside of PHCs and RHs, underlying stock-outs and the limitations in public structures delivering less costly treatment than in private pharmacies. Although it is clear that the mobilisation of financial resources is insufficient to support free healthcare policies, there is a need for further research to determine efficient financing modalities to ensure a regular and adequate supply of medicines in public health facilities. Our study thus provides essential information and hypotheses for potential actions aimed at improving health policies towards the affordability of primary healthcare for children under 5, and for better implementation of universal health coverage in sub-Saharan Africa.

## Acknowledgements

**Children, families, UNITAID and The AIRE Research Study Group: Country investigators:** Ouagadougou, Burkina Faso: S. Yugbaré Ouédraogo (PI), V. M. Sanon Zombré (CoPI), Conakry, Guinea: M. Sama Cherif (CoPI), I. S. Diallo (CoPI), D. F. Kaba, (PI). Bamako, Mali: A. A. Diakité (PI), A. Sidibé, (CoPI). Niamey, Niger: H. Abarry Souleymane (CoPI), F. Tidjani Issagana Dikouma (PI). **Research coordinators & data centers: Inserm U1295, Toulouse 3 University, France:** H. Agbeci (Int Health Economist), L. Catala (Research associate), D. L. Dahourou (Research associate), S. Desmonde (Research associate), E. Gres (PhD Student), G. B. Hedible (Int research project manager), V. Leroy (research coordinator), L. Peters Bokol (Int clinical research monitor), J. Tavarez (Research project assistant), Z. Zair (Statistician, Data scientist). **CEPED, IRD, Paris, France:** S. Louart (process manager), V. Ridde (process coordination). **Inserm U1137, Paris, France:** A. Cousien (Research associate). **Inserm U1219**, EMR271 IRD, **Bordeaux University, France:** R. Becquet (Research associate), V. Briand (Research associate), V. Journot (Research associate). **PACCI, CHU Treichville, Abidjan, Côte d’Ivoire :** S. Lenaud (Int data manager), C. N’Chot (Research associate), B. Seri (Supervisor IT), C. Yao (data manager supervisor). **Consortium NGOs partners: Alima-HQ (consortium lead), Dakar, Sénégal:** G. Anago (Int Monitoring Evaluation Accountability And Learning Officer), D. Badiane (Supply chain manager), M. Kinda (Director), D. Neboua (Medical officer), P. S. Dia (Supply chain manager), S. Shepherd (referent NGO), N. di Mauro (Operations support officer), G. Noël (Knowledge broker), K. Nyoka (Communication and advocacy officer), W. Taokreo (Finance manager), O. B. Coulidiati Lompo (Finance manager), M. Vignon (Project Manager). **Alima, Conakry, Guinea:** P. Aba (clinical supervisor), N. Diallo (clinical supervisor), M. Ngaradoum (Medical Team Leader), S. Léno (data collector), A. T. Sow (data collector), A. Baldé (data collector), A. Soumah (data collector), B. Baldé (data collector), F. Bah (data collector), K. C. Millimouno (data collector), M. Haba (data collector), M. Bah (data collector), M. Soumah (data collector), M. Guilavogui (data collector), M. N. Sylla (data collector), S. Diallo (data collector), S. F. Dounfangadouno (data collector), T. I. Bah (data collector), S. Sani (data collector), C. Gnongoue (Monitoring Evaluation Accountability And Learning Officer), S. Gaye (Monitoring Evaluation Accountability And Learning Officer), J. P. Y. Guilavogui (Clinical Research Assistant), A. O. Touré (Country health economist), J. S. Kolié (Country clinical research monitor), A. S. Savadogo (country project manager). **Alima, Bamako, Mali:** F. Sangala (Medical Team Leader), M. Traore (Clinical supervisor), T. Konare (Clinical supervisor), A. Coulibaly (Country health economist), A. Keita (data collector), D. Diarra (data collector), H. Traoré (data collector), I. Sangaré (data collector), I. Koné (data collector), M. Traoré (data collector), S. Diarra (data collector), V. Opoue (Monitoring Evaluation Accountability And Learning Officer), F. K. Keita (medical coordinator), M. Dougabka (Clinical research assistant then Monitoring Evaluation Accountability And Learning Officer), B. Dembélé (data collector then Clinical research assistant), M. S. Doumbia (country health economist), G. D. Kargougou (country clinical research monitor), S. Keita (country project manager). **Solthis-HQ, Paris:** S. Bouille (NGO referent), S. Calmettes (NGO referent), F. Lamontagne (NGO referent). **Solthis, Niamey:** K. H. Harouna (clinical supervisor), B. Moutari (clinical supervisor), I. Issaka (clinical supervisor), S. O. Assoumane (clinical supervisor), S. Dioiri (Medical Team Leader), M. Sidi (data collector), K. Sani Alio (Country supply chain officer), S. Amina (data collector), R. Agbokou (Clinical research assistant), M. G. Hamidou (Clinical Research Assistant), S. M. Sani (Country health economist), A. Mahamane, Aboubacar Abdou (data collector), B. Ousmane (data collector), I Kabirou (data collector), I. Mahaman (data collector), I Mamoudou (data collector), M. Baguido (data collector), R. Abdoul (data collector), A. Sahabi (data collector), F. Seini (data collector), Z. Hamani (data collector), L-Y B Meda (Country clinical research monitor), Mactar Niome (country project manager), X. Toviho (Monitoring Evaluation Accountability And Learning Officer), I. Sanouna (Monitoring Evaluation Accountability And Learning Officer), P. Kouam (program officer). **Terre des hommes-HQ, Lausanne:** S. Busière (NGO referent), F. Triclin (NGO referent). **Terre des hommes, BF:** A. Hema (country project manager), M. Bayala (IeDA IT), L. Tapsoba (Monitoring Evaluation Accountability And Learning Officer), J. B. Yaro (Clinical reearch assistant), S. Sougue (Clinical reearch assistant), R. Bakyono (Country health economist), A. G. Sawadogo (Country clinical research monitor), A. Soumah (data collector), Y. A. Lompo (data collector), B. Malgoubri (data collector), F. Douamba (data collector), G. Sore (data collector), L. Wangraoua (data collector), S. Yamponi (data collector), S. I. Bayala (data collector), S. Tiegna (data collector), S. Kam (data collector), S. Yoda (data collector), M. Karantao (data collector), D. F. Barry (Clinical supervisor), O. Sanou (clinical supervisor), N. Nacoulma (Medical Team Leader), N. Semde (clinical supervisor), I. Ouattara (Clinical supervisor), F. Wango (clinical supervisor), Z. Gneissien (clinical supervisor), H. Congo (clinical supervisor). **Terre des hommes, Mali:** Y. Diarra (clinical supervisor), B. Ouattara (clinical supervisor), A. Maiga (data collector), F. Diabate (data collector), O. Goita (data collector), S. Gana (data collector), S. Diallo (data collector), S. Sylla (data collector), D. Coulibaly (Tdh project manager), N. Sakho (NGO referent). **Country SHS team: Burkina Faso:** K. Kadio (consultant and research associate), J. Yougbaré (data collector), D. Zongo (data collector), S. Tougouma (data collector), A. Dicko (data collector), Z. Nanema (data collector), I. Balima (data collector), A. Ouedraogo (data collector), A. Ouattara (data collector), S. E. Coulibaly (data collector). **Guinea:** H. Baldé (consultant and research associate), L. Barry (data collector), E. Duparc Haba (data collector). **Mali:** A. Coulibaly (consultant and research associate), T. Sidibe (data collector), Y. Sangare (data collector), B. Traore (data collector), Y. Diarra (data collector). **Niger:** A. E. Dagobi (consultant and research associate), S. Salifou (data collector), B. Gana Moustapha Chétima (data collector), I. H. Abdou (data collector)

## Authors’ contributions

HA, SD, GBH and VL conceived and designed the study. HA, GBH, ZZ and LPB, RB, AO, AC carried out data collection and data curation. HA, ZZ, analysed the data with SD and VL supervision. HA wrote the first draft of the manuscript. All authors were involved in data interpretation and review of the final manuscript. VL is the guarantor to submit the manuscript.

## Data Availability Statement

The datasets generated and analysed during the current study are not publicly available. Access to processed deidentified participant data will be made available to any third Party after the publication of the main AIRE results stated in the Pan African Clinical Trial Registry Study statement (PACTR202206525204526, registered on 06/15/2022), upon a motivated request (concept sheet), and after the written consent of the AIRE research coordinator (Valeriane Leroy, Inserm U1295 Toulouse, France,) obtained after the approval of the AIRE publication committee, if still active.

## Appendix

## Appendix 1

**Table:** Comparison of children selected for the cost study versus unselected children according to disease severity and country in the AIRE project, June 2021 to May 2022.

|  | **Burkina Faso (N=4,834)** |  |  |  |  |  |
| --- | --- | --- | --- | --- | --- | --- |
|  | Non-severe cases (N=4 592) |  |  | Severe cases (N=242) |  |  |
|  | No cost sample<br>(n=4 353) | Cost sample<br>(n=239) | *p-value* | No cost sample<br>(n=96) | Cost sample<br>(n=146) | *p-value* |
| **Child's age:** median age in month | 19 | 18 | 0.580 | 24 | 26 | 0.832 |
| **Child's sex:** Male (%) | 52 | 55 | 0.332 | 52 | 60 | 0.249 |
| **Education level of head household:** Primary level and up (%) | 82 | 80 | 0.426 | 15 | 16 | 0.805 |
| **Income-generating activity of the main child caregiver:** Yes (%) | 20 | 26 | 0.032 | 17 | 16 | 0.963 |
| **Consultation delay from from the onset symptoms:** less than 2 days (%) | 45 | 44 | 0.575 | 38 | 45 | 0.235 |
| **Transport modalities:** Foot/Cart/Bike (%) | 84 | 81 | 0.176 | 69 | 59 | 0.121 |
|  | **Guinea (N=1,406)** |  |  |  |  |  |
|  | Non-severe cases (N=778) |  |  | Severe cases (N=628) |  |  |
|  | No cost sample<br>(n=547) | Cost sample<br>(n=231) | *p-value* | No cost sample<br>(n=417) | Cost sample<br>(n=211) | *p-value* |
| **Child's age:** median age in month | 15 | 19 | 0.125 | 24 | 24 | 0.793 |
| **Child's sex:** Male (%) | 52 | 49 | 0.574 | 49 | 54 | 0.273 |
| **Education level of head household:** Primary level and up (%) | 49 | 52 | 0.452 | 54 | 47 | 0.120 |
| **Income-generating activity of the main child caregiver:** Yes (%) | 50 | 55 | 0.297 | 51 | 51 | 0.979 |
| **Consultation delay from from the onset symptoms:** less than 2 days (%) | 15 | 16 | 0.731 | 10 | 14 | 0.116 |
| **Transport modalities:** Foot/Cart/Bike (%) | 30 | 37 | 0.037 | 27 | 34 | 0.088 |

|  | **Mali (N=4,259)** |  |  |  |  |  |
| --- | --- | --- | --- | --- | --- | --- |
|  | No severe cases (N=3 455) |  |  | Severe cases (N=804) |  |  |
|  | No cost sample<br>(n=3 217) | Cost sample<br>(n=238) | *p-value* | No cost sample<br>(n=602) | Cost sample<br>(n=202) | *p-value* |
| **Child's age:** median age in month | 18 | 17 | 0.305 | 24 | 20 | 0.256 |
| **Child's sex:** Male (%) | 54 | 58 | 0.241 | 52 | 52 | 0.965 |
| **Education level of head household:** Primary level and up (%) | 69 | 64 | 0.120 | 42 | 31 | 0.004 |
| **Income-generating activity of the main child caregiver:** Yes (%) | 24 | 32 | 0.007 | 19 | 28 | 0.006 |
| **Consultation delay from from the onset symptoms:** less than 2 days (%) | 30 | 33 | 0.385 | 25 | 28 | 0.329 |
| **Transport modalities:** Foot/Cart/Bike (%) | 26 | 28 | 0.512 | 11 | 20 | 0.001 |
|  | **Niger (N=5,337)** |  |  |  |  |  |
|  | Non-severe cases (N=5 013) |  |  | Severe cases (N=324) |  |  |
|  | No cost sample<br>(n=4 781) | Cost sample<br>(n=232) | *p-value* | No cost sample<br>(n=138) | Cost sample<br>(n=186) | *p-value* |
| **Child's age:** median age in month | 15 | 13 | 0.195 | 10 | 16 | 0.000 |
| **Child's sex:** Male (%) | 53 | 53 | 0.845 | 49 | 54 | 0.371 |
| **Education level of head household:** Primary level and up (%) | 51 | 57 | 0.060 | 41 | 36 | 0.003 |
| **Income-generating activity of the main child caregiver:** Yes (%) | 21 | 25 | 0.224 | 7 | 14 | 0.057 |
| **Consultation delay from from the onset symptoms:** less than 2 days (%) | 48 | 45 | 0.314 | 28 | 18 | 0.035 |
| **Transport modalities:** Foot/Cart/Bike (%) | 81 | 80 | 0.667 | 53 | 63 | 0.056 |

## Appendix 2

**Figure 1:**
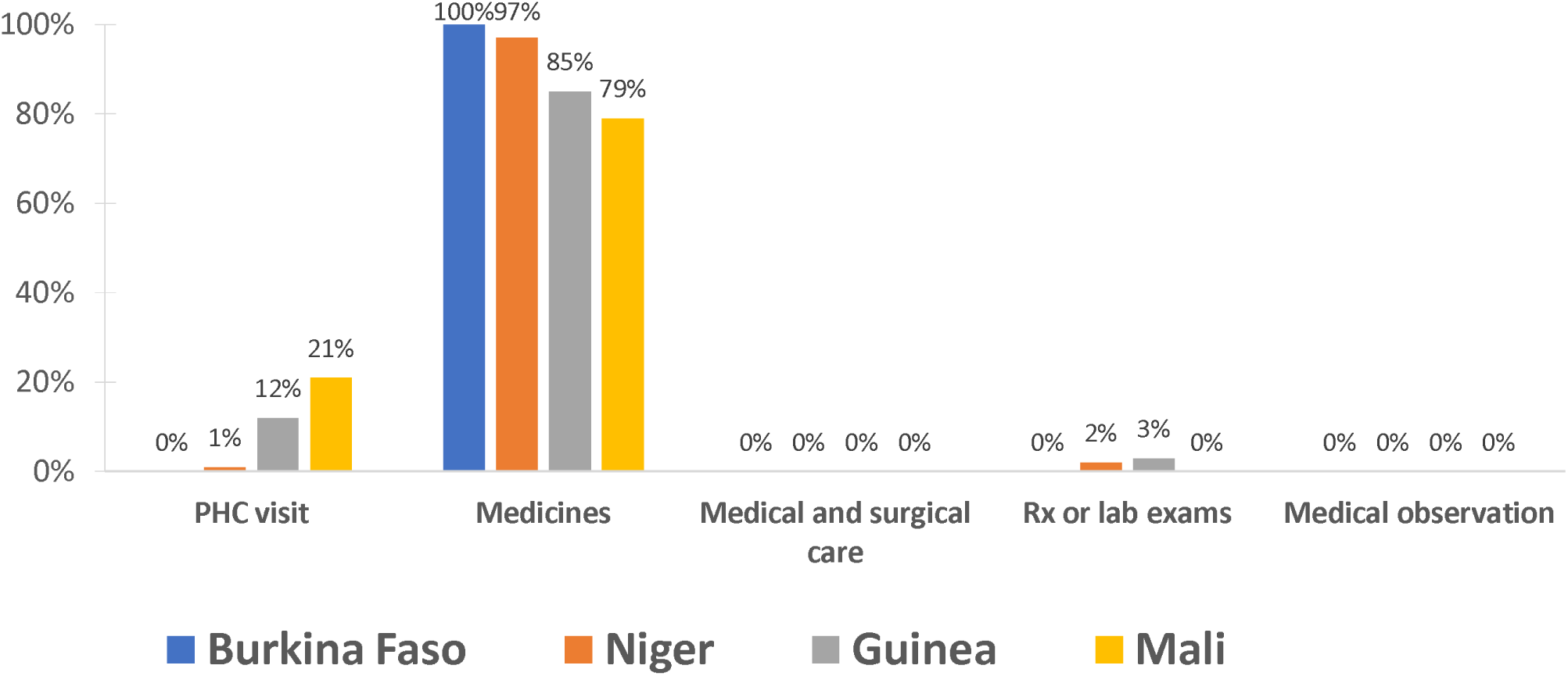
Distribution of different types of medical direct costs for non-severe cases according to country.

**Figure 2:**
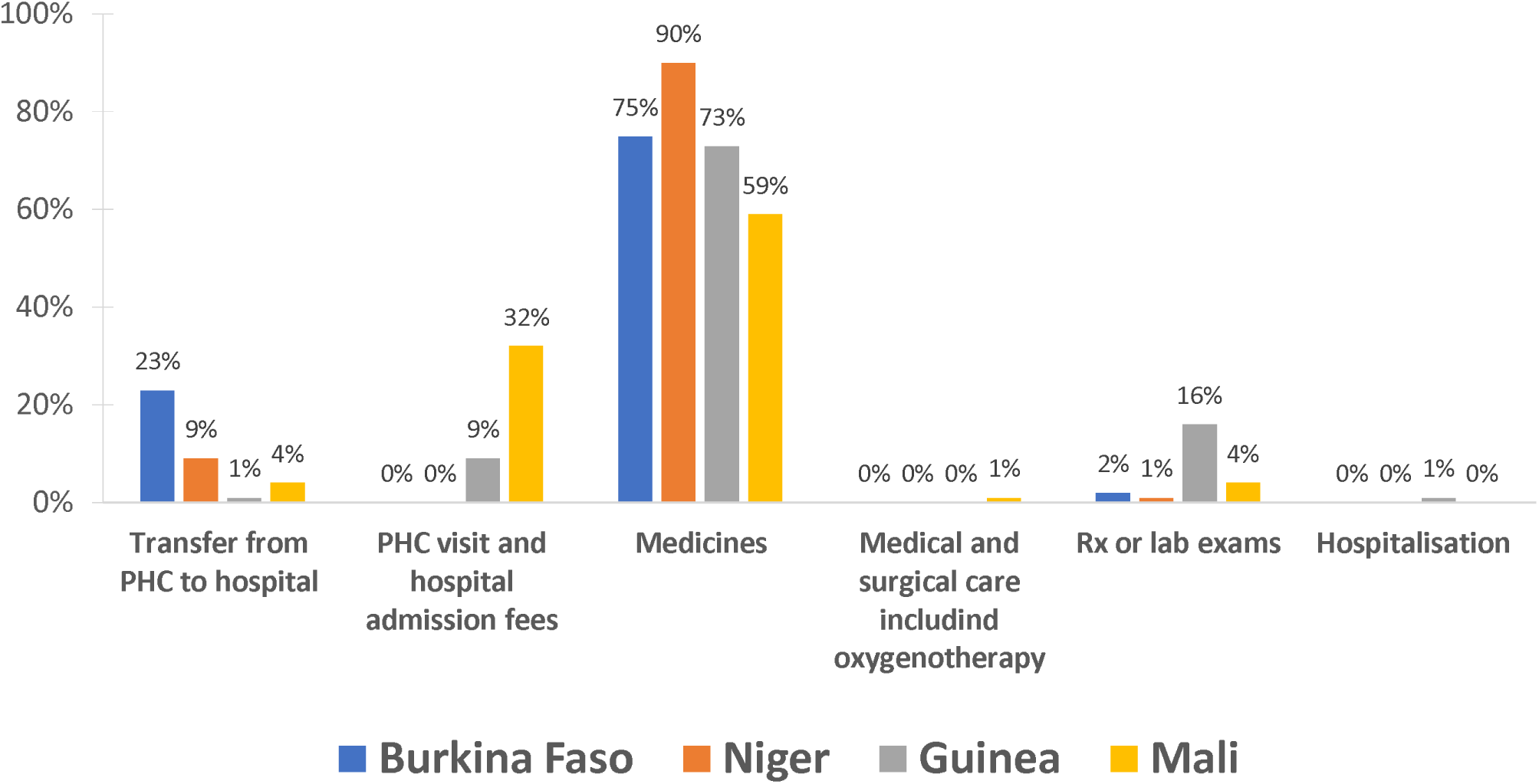
Distribution of different types of medical direct costs for severe cases according to country.

**Table:** Number (percentage), mean (standard deviation), minimum and maximum of medicines cost per episode of illness of a child under the age of 5 among children whose households incurred expenses, according to severity and country.

|  | Number | Percentage | Mean | Standard deviation | Min | Max |
| --- | --- | --- | --- | --- | --- | --- |
| **Non-severe cases** |  |  |  |  |  |  |
| **Burkina Faso (N=239)** | 65 | 27.2% | 3.4 | 2.8 | 0.6 | 17.0 |
| **Guinea (N=231)** | 226 | 97.8% | 6.5 | 3.2 | 0.0 | 16.8 |
| **Mali (N=238)** | 237 | 99.6% | 4.6 | 2.9 | 0.0 | 25.2 |
| **Niger (N=232)** | 201 | 86.6% | 4.5 | 2.6 | 0.0 | 13.3 |
| **Severe cases** |  |  |  |  |  |  |
| **Burkina Faso (N=146)** | 83 | 56.8% | 7.4 | 8.5 | 0.0 | 44.9 |
| **Guinea (N=211)** | 171 | 81.0% | 10.7 | 9.0 | 0.0 | 52.9 |
| **Mali (N=202)** | 193 | 95.5% | 8.1 | 8.9 | 0.0 | 56.4 |
| **Niger (N=186)** | 173 | 93.0% | 17.5 | 13.5 | 0.0 | 60.5 |

## Appendix 3

**Table:** Number (percentage), mean (standard deviation), minimum and maximum transfer costs from primary healthcare centre to referral hospital among children with a severe case for whom households incurred transfer costs according to the country.

| Countries | Severe cases | Referred to HR accepted n (%) | Referred cases with household transfer expenditure | Mean of household transfer expenditure | Standard deviation | Min | Max |
| --- | --- | --- | --- | --- | --- | --- | --- |
| Burkina Faso | 146 | 43 (29) | 43 | 3 | 1.6 | 0.5 | 6.5 |
| Guinea | 211 | 19 (9) | 19 | 2.7 | 3.5 | 0.3 | 13.2 |
| Mali | 202 | 34 (17) | 31 | 11.8 | 27.3 | 1.4 | 99.5 |
| Niger | 186 | 136 (73) | 70 | 6.1 | 6.8 | 0.5 | 22.5 |

## Appendix 4

**Table:** Number (percentage), mean (standard deviation), minimum and maximum per episode of illness for a child whose households incurred expenses, according to severity, type of cost and country for children under-5 enrolled in the AIRE cost study.

|  | Total exemption user fees policy |  |  |  |  |  | Partial exemption user fees policy |  |  |  |  |  |
| --- | --- | --- | --- | --- | --- | --- | --- | --- | --- | --- | --- | --- |
|  | Burkina Faso |  |  | Niger |  |  | Guinea |  |  | Mali |  |  |
|  | Non-severe cases (N=239) | severe cases (N=146) | All cases (N=385) | Non-severe cases (N=232) | Severe cases (N=186) | All cases (N=418) | Non-severe cases (N=231) | Severe cases (N=211) | All cases (N=442) | Non-severe cases (N=238) | Severe cases (N=202) | All cases (N=440) |
| **Medical direct cost** |  |  |  |  |  |  |  |  |  |  |  |  |
| Number (%) | 65 (27.2) | 83 (56.8) | 148 (38.4) | 201 (86.6) | 173 (93.0) | 374 (89.4) | 226 (97.8) | 171 (81.0) | 397 (89.8) | 237 (99.6) | 193 (95.5) | 430 (97.7) |
| Mean (SD) | 3.4 (2.8) | 9.9 (12.1) | 7.0 (9.7) | 4.6 (2.7) | 20.2 (16.6) | 11.8 (13.8) | 7.5 (3.6) | 14.9 (14.5) | 10.6 (10.6) | 5.5 (2.9) | 12.8 (18.6) | 8.8 (13.2) |
| Min | 0.6 | 0.5 | 0.5 | 0.5 | 1.8 | 0.5 | 0.5 | 0.4 | 0.4 | 0.9 | 0.4 | 0.4 |
| Max | 16.9 | 74.8 | 74.8 | 13.5 | 78.5 | 78.5 | 20.3 | 73.2 | 73.2 | 26.1 | 133.8 | 133.8 |
| **Non-medical direct cost** |  |  |  |  |  |  |  |  |  |  |  |  |
| Number (%) | 9 (3.8) | 84 (57.5) | 93 (24.1) | 4 (2.7) | 136 (73.1) | 140 (33.5) | 18 (7.8) | 147 (69.7) | 165 (37.3) | 0 | 171 (84.6) | 171 (38.9) |
| Mean (SD) | 1.2 (0.4) | 3.7 (6.9) | 3.5 (6.6) | 1.5 (0.6) | 3.2 (3.3) | 3.2 (3.2) | 1.9 (1.0) | 1.8 (1.2) | 1.8 (1.2) | - | 2.6 (7.8) | 2.6 (7.8) |
| Min | 0.5 | 0.3 | 0.3 | 0.7 | 0.4 | 0.4 | 0.3 | 0.4 | 0.3 | - | 0.3 | 0.3 |
| Max | 1.6 | 57.7 | 57.7 | 2.2 | 20.9 | 20.9 | 4.3 | 6.6 | 6.6 | - | 101.0 | 101.0 |
| **Indirect cost** |  |  |  |  |  |  |  |  |  |  |  |  |
| Number (%) | 16 (6.7) | 58 (39.7) | 74 (19.2) | 0 | 5 (2.7) | 5 (1.2) | 40 (17.3) | 92 (43.6) | 132 (29.9) | 30 (12.6) | 103 (50.9) | 133 (30.2) |
| Mean (SD) | 3.0 (1.3) | 8.0 (6.2) | 6.9 (5.9) | - | 29.7 (22.9) | 29.7 (22.9) | 6.1 (5.4) | 7.9 (5.7) | 7.3 (5.6) | 4.8 (1.1) | 12.7 (13.4) | 10.9 (12.2) |
| Min | 1.8 | 1.8 | 1.8 | - | 4.5 | 4.5 | 3.6 | 3.6 | 3.5 | 4.5 | 4.5 | 4.5 |
| Max | 5.4 | 21.6 | 21.6 | - | 54.1 | 54.1 | 28.5 | 32.0 | 32.0 | 9 | 54.1 | 54.1 |

## Appendix 5

**Table:**
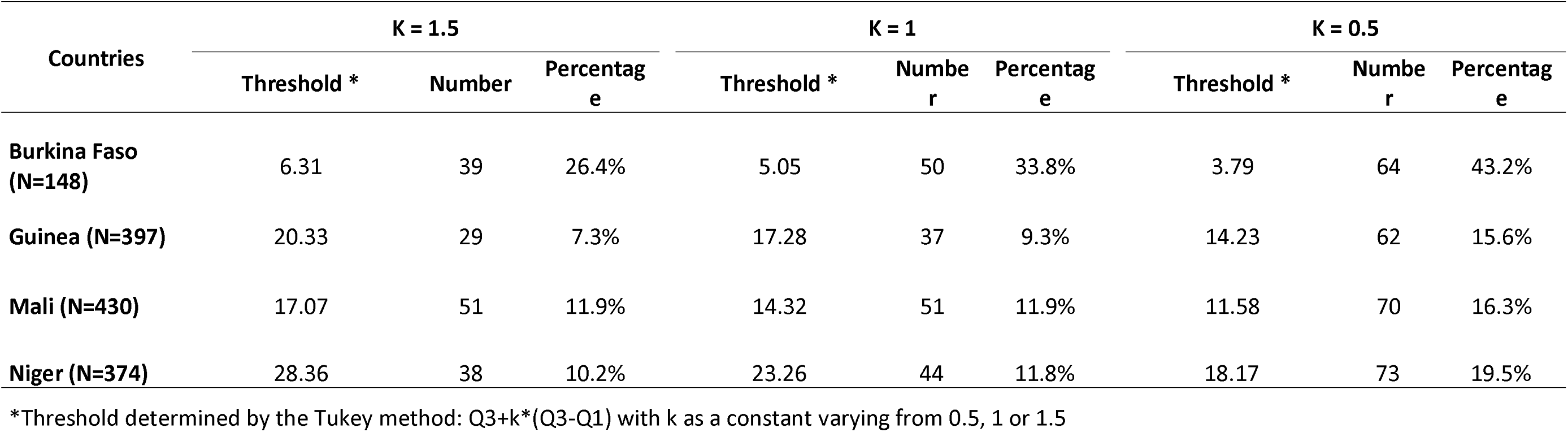
Number and percentage of households with excessive expenditure among households with direct payment for the care of children under 5 according to country.

| Countries | K = 1.5 |  |  | K = 1 |  |  | K = 0.5 |  |  |
| --- | --- | --- | --- | --- | --- | --- | --- | --- | --- |
|  | Threshold * | Number | Percentage | Threshold * | Number | Percentage | Threshold * | Number | Percentage |
| **Burkina Faso (N=148)** | 6.31 | 39 | 26.4% | 5.05 | 50 | 33.8% | 3.79 | 64 | 43.2% |
| **Guinea (N=397)** | 20.33 | 29 | 7.3% | 17.28 | 37 | 9.3% | 14.23 | 62 | 15.6% |
| **Mali (N=430)** | 17.07 | 51 | 11.9% | 14.32 | 51 | 11.9% | 11.58 | 70 | 16.3% |
| **Niger (N=374)** | 28.36 | 38 | 10.2% | 23.26 | 44 | 11.8% | 18.17 | 73 | 19.5% |

\*Threshold determined by the Tukey method: $Q3 + k \cdot (Q3 - Q1)$ with k as a constant varying from 0.5, 1 or 1.5

**Appendix 6:** CHEERS 2022 Checklist.

|  | Item | Guidance for Reporting | Reported in section |
| --- | --- | --- | --- |
| **TITLE** |  |  |  |
| Title | 1 | Identify the study as an economic evaluation and specify the interventions being compared. | Yes |
| **ABSTRACT** |  |  |  |
| Abstract | 2 | Provide a structured summary that highlights context, key methods, results and alternative analyses. | Yes |
| **INTRODUCTION** |  |  |  |
| Background and objectives | 3 | Give the context for the study, the study question and its practical relevance for decision making in policy or practice. | Yes |
| **METHODS** |  |  |  |
| Health economic analysis plan | 4 | Indicate whether a health economic analysis plan was developed and where available. | Not developed/available |
| Study population | 5 | Describe characteristics of the study population (such as age range, demographics, socioeconomic, or clinical characteristics). | Yes |
| Setting and location | 6 | Provide relevant contextual information that may influence findings. | Yes |
| Comparators | 7 | Describe the interventions or strategies being compared and why chosen. | Not relevant |
| Perspective | 8 | State the perspective(s) adopted by the study and why chosen. | Yes |
| Time horizon | 9 | State the time horizon for the study and why appropriate. | Yes |
| Discount rate | 10 | Report the discount rate(s) and reason chosen. | Not relevant |
| Selection of outcomes | 11 | Describe what outcomes were used as the measure(s) of benefit(s) and harm(s). | Not relevant |
| Measurement of outcomes | 12 | Describe how outcomes used to capture benefit(s) and harm(s) were measured. | Not relevant |
| Valuation of outcomes | 13 | Describe the population and methods used to measure and value outcomes. | Not relevant |
| Measurement and valuation of resources and costs | 14 | Describe how costs were valued. | Yes |
| Currency, price date, and conversion | 15 | Report the dates of the estimated resource quantities and unit costs, plus the currency and year of conversion. | Yes |
| Rationale and description of model | 16 | If modelling is used, describe in detail and why used. Report if the model is publicly available and where it can be accessed. | Not relevant |
| Analytics and assumptions | 17 | Describe any methods for analysing or statistically transforming data, any extrapolation methods, and approaches for validating any model used. | Yes |
| Characterizing heterogeneity | 18 | Describe any methods used for estimating how the results of the study vary for sub-groups. | Yes |
| Characterizing distributional effects | 19 | Describe how impacts are distributed across different individuals or adjustments made to reflect priority populations. | Not relevant |
| Characterizing uncertainty | 20 | Describe methods to characterize any sources of uncertainty in the analysis. | Not relevant |
| Approach to engagement with patients and others affected by the study | 21 | Describe any approaches to engage patients or service recipients, the general public, communities, or stakeholders (e.g., clinicians or payers) in the design of the study. | Yes |
| **RESULTS** |  |  |  |
| Study parameters | 22 | Report all analytic inputs (e.g., values, ranges, references) including uncertainty or distributional assumptions. | Yes |
| Summary of main results | 23 | Report the mean values for the main categories of costs and outcomes of interest and summarise them in the most appropriate overall measure. | Yes |
| Effect of uncertainty | 24 | Describe how uncertainty about analytic judgments, inputs, or projections affect findings. Report the effect of choice of discount rate and time horizon, if applicable. | Not applicable |
| Effect of engagement with patients and others affected by the study | 25 | Report on any difference patient/service recipient, general public, community, or stakeholder involvement made to the approach or findings of the study | Yes |
| **DISCUSSION** |  |  |  |
| Study findings, limitations, generalizability, and current knowledge | 26 | Report key findings, limitations, ethical or equity considerations not captured, and how these could impact patients, policy, or practice. | Yes |
| **OTHER RELEVANT INFORMATION** |  |  |  |
| Source of funding | 27 | Describe how the study was funded and any role of the funder in the identification, design, conduct, and reporting of the analysis | Yes |
| Conflicts of interest | 28 | Report authors conflicts of interest according to journal or International Committee of Medical Journal Editors requirements. | Yes |

Husereau D, Drummond M, Augustovski F, de Bekker-Grob E, Briggs AH, Carswell C, Caulley L, Chaiyakunapruk N, Greenberg D, Loder E, Mauskopf J, Mullins CD, Petrou S, Pwu RF, Staniszewska S; CHEERS 2022 ISPOR Good Research Practices Task Force. Consolidated Health Economic Evaluation Reporting Standards 2022 (CHEERS 2022) Statement: Updated Reporting Guidance for Health Economic Evaluations. *BMJ*. 2022;376:e067975. The checklist is Open Access distributed in accordance with the terms of the Creative Commons Attribution (CC BY 4.0) license, which permits others to distribute, remix, adapt and build upon this work, for commercial use, provided the original work is properly cited. See: http://creativecommons.org/licenses/by/4.0/.

